# Tumor Resectability and Pathologic Response After Neoadjuvant Long-Course Chemoradiotherapy for Locally Advanced Rectal Cancer in a Resource-Limited Setting

**DOI:** 10.64898/2026.03.25.26349327

**Authors:** Samrawit S. Halake, Hawi F. Bedada, Tariku M. Desalegn, Tsige B. Feyisa, Kibrekidusan A. Tsige, Edom S. Woldetsadik, Eva J. Kantelhardt

**Affiliations:** Department of Clinical Oncology, Tikur Anbessa Specialized Hospital, School of Medicine, College of Health Science, Addis Ababa University, Addis Ababa, Ethiopia; Department of Oncology, Kampala International University, Ishaka, Uganda; Oncology Unit, Saint Paul’s Hospital Millennium Medical College, Addis Ababa, Ethiopia; Armauer Hansen Research Institute, Addis Ababa, Ethiopia; Department of Gynecology, Martin-Luther-University, Halle (Saale), Germany

**Keywords:** Locally advanced rectal cancer, long-course chemoradiotherapy, tumor resectability, pathologic response

## Abstract

**Purpose:** In resource-limited settings, locally advanced rectal cancer (LARC) often presents at advanced stages. Long-course chemoradiotherapy (LCCRT) remains a cornerstone of neoadjuvant therapy, yet outcome data from such settings remain limited. This study assessed tumor resectability, pathologic response, and factors associated with resectability following neoadjuvant LCCRT at Ethiopia’s largest tertiary oncology center.

**Methods:** A retrospective cohort study was conducted among patients with stage II–III rectal adenocarcinoma (cT3–4 and/or cN+) who completed neoadjuvant LCCRT at Tikur Anbessa Specialized Hospital between 2018 and 2023. Tumor resectability was determined by multidisciplinary team (MDT) assessment. Multivariable logistic regression was used to identify factors associated with post-LCCRT resectability, adjusting for initial T stage, circumferential resection margin (CRM) status, histologic subtype, radiotherapy technique, and neoadjuvant regimen.

**Results:** Among 58 eligible patients (median age 45 years; 62% male), 62% had cT4 tumors, 53% had cN2 disease, and 84.5% had involved CRM. The median diagnosis-to-LCCRT interval was 64 weeks (interquartile range [IQR], 37–82). After LCCRT, 27 patients (46.6%) were deemed resectable by MDT assessment; 19 patients (32.8%) ultimately underwent curative-intent surgery (median interval from LCCRT to surgery, 10 weeks; IQR, 7–15). Initial cT3 stage was associated with higher odds of resectability (adjusted odds ratio [AOR], 6.2; 95% CI, 1.06–36.37), whereas receipt of total neoadjuvant therapy was associated with lower odds (AOR, 0.10; 95% CI, 0.02– 0.49). No pathologic complete responses were observed.

**Conclusion:** In this cohort characterized by advanced disease at presentation and treatment delays, neoadjuvant LCCRT resulted in low resectability and limited pathologic response. To enhance curative potential, concerted efforts are needed to expedite the timely initiation of radiotherapy, optimize multidisciplinary team assessment, and increase surgical capacity.

**Context:** *Key objective:* How often does neoadjuvant long-course chemoradiotherapy result in tumor resectability and pathologic response among patients with locally advanced rectal cancer treated in a resource-limited setting, and which factors are associated with post-treatment resectability?

*Knowledge generated:* In a cohort with predominantly advanced rectal cancer and prolonged treatment intervals, fewer than half of tumors were considered resectable after long-course chemoradiotherapy, and only one-third of patients underwent curative-intent surgery. No pathologic complete responses were observed, and an earlier T stage was the main factor associated with resectability.

*Relevance:* These findings from a resource-limited setting highlight real-world challenges in managing locally advanced rectal cancer and provide evidence to guide clinical decision-making and multidisciplinary planning. They emphasize the potential benefits of timely radiotherapy access, coordinated multidisciplinary care, and surgical expertise to improve resectability, increase curative resections, and reduce outcome disparities.

## Introduction

Rectal Cancer is the second most common gastrointestinal malignancy worldwide, following Colon Cancer (1). In recent decades, colorectal cancer (CRC) incidence and mortality have increased disproportionately in low- and middle-income countries (LMICs), reflecting changes in population demographics, lifestyle factors, and health system capacity (2-4). In Ethiopia, CRC incidence is rising and currently ranks as the most common cancer among men and the third most common among women (5, 6). Rectal cancer is among the frequently diagnosed and managed malignancies at Tikur Anbessa Specialized Hospital (TASH), the largest oncology center in Ethiopia (7, 8)

Management of nonmetastatic rectal cancer relies on a multimodal approach that includes surgery, radiotherapy, systemic therapy, and, in selected cases, immunotherapy (9, 10). Surgical resection with total mesorectal excision (TME) remains the cornerstone of curative treatment and is critical for achieving optimal local control (11). While early-stage rectal cancers are generally managed with upfront surgery with or without adjuvant therapy, locally advanced rectal cancer (LARC) requires coordinated multimodality treatment (12). Multidisciplinary team (MDT)–based decision-making is recommended for LARC and typically involves medical and radiation oncologists, colorectal surgeons, radiologists, pathologists, and specialized nursing staff. MDT discussions have been associated with improved adherence to evidence-based care and better patient outcomes (13, 14).

According to the 8th edition of the American Joint Committee on Cancer (AJCC) tumor–node–metastasis (TNM) staging system, neoadjuvant therapy is definitively indicated for nonmetastatic rectal cancers staged as clinical (c) T3 or cT4 (15). These represent groups of patients who will ultimately require postoperative radiotherapy if resected initially. Other relative indications for Neoadjuvant therapy include mesorectal involvement or clinical nodal disease in cT1 and cT2 Rectal Cancer (12, 16).

The role of preoperative radiotherapy in LARC was established by landmark randomized trials demonstrating superior local control with neoadjuvant compared with adjuvant treatment (17). Meta-analyses have further shown significantly lower local recurrence rates with preoperative chemoradiotherapy (CRT) relative to postoperative CRT (18). Even though survival is reported to be similar in Neoadjuvant CRT and Adjuvant CRT, increased rates of resectability and pathologic response seen with the Neoadjuvant approach have made it the favored approach in many institutions (19-22), including TASH (23). Additional advantages of the preoperative approach include increased likelihood of sphincter-preserving surgery, improved radiosensitivity of well-oxygenated tissues, reduced small bowel toxicity, and resection of irradiated tissues at the time of surgery (12, 18).

Neoadjuvant treatment can be delivered as Long-course Chemoradiotherapy (LCCRT), Short-course Radiotherapy (SCRT), or more recently Total Neoadjuvant Treatment (TNT) which entails delivering both radiation and chemotherapy before definitive surgery (19-22). International Guidelines, including the European Society for Medical Oncology (ESMO) and National Comprehensive Cancer Network (NCCN), have endorsed these approaches, thus making Neoadjuvant treatment a standard of care for LARC patients (12, 16).

Despite these advances, implementation of guideline-based neoadjuvant strategies remains challenging in resource-limited settings. Among patients who complete standard neoadjuvant treatment, data describing tumor resectability and pathologic response are limited. This study evaluates resectability rates and pathologic response among patients with LARC treated with neoadjuvant LCCRT at TASH, providing insight into treatment outcomes and influencing factors within a resource-constrained tertiary care setting.

## Method

### Study Design and Setting

This was a retrospective institutional-based cohort study conducted at TASH, Addis Ababa University, Ethiopia. TASH is the country’s largest tertiary oncology referral center, providing comprehensive cancer care, including chemotherapy, radiotherapy, surgery, and palliative services for patients referred nationwide.

### Study Population

The study included adult patients with histologically confirmed rectal adenocarcinoma who received neoadjuvant long-course chemoradiotherapy (LCCRT) at the adult oncology department of TASH between October 2018 and October 2023. Patients were eligible if they had locally advanced rectal cancer (LARC) and completed the planned course of neoadjuvant LCCRT followed by post-treatment multidisciplinary team (MDT) assessment. Patients were excluded if medical records lacked key clinical or treatment information (including baseline staging or neoadjuvant treatment details), if radiotherapy was not completed, or if post-treatment MDT assessment was not documented.

### Patient Selection

During the study period, 87 patients with LARC were initiated on neoadjuvant LCCRT following MDT discussion or decision by a senior oncologist. Of these, nine patients were excluded because of incomplete clinical documentation, and 20 patients did not complete the prescribed radiotherapy course. The final analytic cohort consisted of 58 patients.

### Treatment and Multidisciplinary Assessment

Neoadjuvant treatment consisted of long-course pelvic radiotherapy delivered with concurrent oral capecitabine. Decisions regarding treatment strategy, including the use of LCCRT alone or the addition of preoperative chemotherapy as part of a total neoadjuvant treatment (TNT) approach, were made through MDT discussions involving at least a colorectal surgeon, body-imaging subspeciality radiologist, and clinical oncologist. Tumor resectability following neoadjuvant treatment was determined by MDT consensus based on post-treatment imaging.

### Study Variables

The primary outcomes were tumor resectability following neoadjuvant therapy and pathologic response among patients who underwent surgical resection. Independent variables included sociodemographic characteristics (age, sex, residence, and documented habits), clinical variables (Eastern Cooperative Oncology Group [ECOG] performance status, tumor location, clinical T and N stage, histologic subtype, and tumor grade), and treatment-related variables (neoadjuvant treatment modality, interval from diagnosis to initiation of radiotherapy, interval from completion of neoadjuvant therapy to surgery, and receipt of additional neoadjuvant chemotherapy).

### Operational definitions

Locally advanced rectal cancer was defined as rectal carcinoma staged as cT3, cT4, or node-positive disease according to the American Joint Committee on Cancer (AJCC) 8th edition staging system (15). Performance status was assessed using the ECOG scale (24). Tumor location was classified as low rectal (<5 cm from the anal verge), mid-rectal (5–10 cm), or high rectal (10–15 cm), based on MRI or Colonoscopic documentation. Tumor size was defined as the mean of two maximal dimensions measured on pelvic MRI. Circumferential resection margin (CRM) status was determined on MRI as the minimum distance between tumor and mesorectal fascia; CRM was considered involved or threatened when this distance was ≤1 mm (15).

Post-neoadjuvant tumor resectability was defined by MDT assessment based on imaging or intraoperative findings. Pathologic response was defined by comparison of pretreatment clinical staging and post-treatment pathologic staging. Complete pathologic response was defined as the absence of viable tumor cells in the resected specimen. Partial pathologic response was defined as downstaging without complete response. No response was defined as unchanged stage, and pathologic upstaging as an increase in pathologic stage relative to pretreatment clinical stage.

### Data collection and statistical analysis

Data were extracted from medical records using a standardized abstraction tool by two oncology residents under supervision. Training was provided before data collection, and a pretest was conducted on approximately 10% of the cohort to refine the abstraction process; these cases were retained in the final analysis due to the limited sample size. Data completeness and consistency were verified by the principal investigator and supervisor.

Data were coded, cleaned, and analyzed using SPSS version 26. Descriptive statistics were used to summarize patient characteristics and treatment variables. Associations between independent variables and tumor resectability were explored using binary logistic regression. Variables with a p value ≤0.25 in bivariable analysis were included in multivariable logistic regression models. Statistical significance was defined as a two-sided p value <0.05, and results were reported as odds ratios with 95% confidence intervals.

### Ethical Considerations

Ethical approval was obtained from the Institutional Review Board of the Department of Clinical Oncology, School of Medicine, Addis Ababa University. Patient confidentiality was maintained by anonymizing data and restricting use to research purposes only.

## Result

### Baseline Demographic and Clinical Characteristics

The median age at diagnosis was 45 years (interquartile range [IQR], 35–60), with 60.3% of patients aged ≤50 years. Slight female predominance was observed (51.7%), and 41.4% of patients resided in Addis Ababa. Most patients had no documented comorbidities (68.9%) and no history of substance use (93.1%). Family history of colorectal cancer was documented in only one patient; however, this variable was not assessed in 72.4% of cases.

The majority of the patients (87.9%) had an ECOG performance status of I, and rectal bleeding was the predominant presenting symptom in 26 (44.8 %) patients. Low rectal tumors predominated, reported in 79.3% by colonoscopy and 87.9% by imaging. Pretreatment tumor size was documented in 44 patients, with a median size of 5.6 cm (range, 1–10 cm). At baseline staging, 62.1% had cT4 disease, and 53.4% had cN2 nodal involvement. CRM was reported in 84.5% following MDT review of pelvic MRI. Adenocarcinoma, not otherwise specified (AC, NOS), accounted for 82.8% of tumors, and 67.2% were well differentiated. (Table 1)

**Table 1:** Baseline Characteristics of Locally Advanced Rectal Cancer Patients at Tikur Anbessa Specialized Hospital, Ethiopia, October 2018 to October 2023. (N=58)

| Characteristics | Category | Frequency | Percentage (%) |
| --- | --- | --- | --- |
| Age in years | $\leq 50$ | 35 | 60.3 |
| | $> 50$ | 23 | 39.7 |
| Sex | Male | 28 | 48.3 |
|  | Female | 30 | 51.7 |
| Residency | Addis Ababa | 24 | 41.4 |
|  | Out of Addis Ababa | 34 | 58.6 |
| Comorbidity | No | 40 | 68.9 |
|  | Yes | 18 | 31.1 |
| **Substance history** | No | 54 | 93.1 |
|  | Chat chewing | 2 | 3.4 |
|  | Not documented | 2 | 3.4 |
| **Family History of CRC** | No | 15 | 25.9 |
|  | Yes | 1 | 1.7 |
|  | Not documented | 42 | 72.4 |
| **ECOG** | I | 51 | 87.9 |
|  | II | 7 | 12.1 |
| **Site based on Colonoscopy** | Low-Rectum | 46 | 79.3 |
|  | Mid-Rectum | 7 | 12.1 |
|  | Upper Rectum | 4 | 6.9 |
|  | Not assessed | 1 | 1.7 |
| **Site based on Imaging** | Low-Rectum | 51 | 87.9 |
|  | Mid-Rectum | 5 | 8.6 |
|  | Upper-Rectum | 2 | 3.5 |
| **Initial T stage** | cT3 | 22 | 37.9 |
|  | cT4 | 36 | 62.1 |
| **Initial N stage** | cN0 | 12 | 20.7 |
|  | cN1 | 15 | 25.9 |
|  | cN2 | 31 | 53.4 |
| **CRM** | Involved | 49 | 84.5 |
|  | Clear | 9 | 15.5 |
| **Histologic subtypes** | AC, NOS | 48 | 82.8 |
|  | Signet Ring Cell | 6 | 10.3 |
|  | Mucinous Carcinoma | 4 | 6.9 |
| **Grade of tumor** | Grade I | 39 | 67.2 |
|  | Grade II | 11 | 19 |
|  | Grade III | 8 | 13.8 |

### Treatment Characteristics

Nearly all patients (98.3%) were discussed at MDT. Initial MDT recommendations favored TNT followed by surgery in 69.0% of cases, while the remainder were planned for LCCRT followed by surgery and adjuvant chemotherapy. Pretreatment diversion colostomy was required in 36.2% of patients.

Neoadjuvant chemotherapy was administered to 25 patients, most commonly using FOLFOX (n=16) or CAPEOX (n=9). Among those receiving chemotherapy, 84% completed six cycles. All patients received concurrent capecitabine during radiotherapy. Two-dimensional radiotherapy (2D-RT) was used in 67.2% of patients, while 32.8% received three-dimensional conformal radiotherapy (3D-CRT). Prescribed radiation doses ranged from 45 to 60 Gy, and the median overall radiotherapy treatment duration was 6.5 weeks (range, 5–13 weeks), with prolongation primarily due to machine-related interruptions.

Following completion of neoadjuvant therapy, 19 patients (32.8%) underwent curative-intent surgery, 32 (55.2%) were managed with palliative treatment, and 7 (12.1%) were placed on clinical follow-up. Among surgical patients, abdominoperineal resection (APR) was performed in 16 (84.2%) and low anterior resection (LAR) in 3 (15.8%). In Table 2, percentages for surgery type are presented using the full cohort (N=58) as the denominator; proportions among surgical patients are described in the text.

**Table 2:** Treatment-related Characteristics of Locally Advanced Rectal Cancer Patients at Tikur Anbessa Specialized Hospital, Ethiopia, October 2018 to October 2023. (N=58)

| Characteristics | Category | Frequency | Percentage (%) |
| --- | --- | --- | --- |
| **Case Discussed at MDT** | Yes | 57 | 98.3 |
|  | No | 1 | 1.7 |
| **Pretreatment diversion colostomy required** | Yes | 21 | 36.2 |
|  | No | 37 | 63.8 |
| **Treatment decision at MDT** | TNT ► Surgery | 40 | 69.0 |
|  | LCCRT ► Surgery ► Chemotherapy | 18 | 31.0 |
| **Delivered treatment** | LCCRT ► Surgery ► Chemotherapy | 19 | 32.8 |
|  | LCCRT ► Chemotherapy | 25 | 43.1 |
|  | LCCRT only | 14 | 24.1 |
| **Neoadjuvant Chemotherapy** | FOLFOX 4 | 16 | 27.6 |
|  | CAPEOX | 9 | 15.5 |
|  | Not administered | 33 | 56.9 |
| **Number of Neoadjuvant chemotherapy cycles given** | 4 | 1 | 1.7 |
|  | 6 | 21 | 36.2 |
|  | 8 | 3 | 5.2 |
|  | Not administered | 33 | 56.9 |
| **Radiotherapy Technique used** | 2D RT | 39 | 67.2 |
|  | 3D CRT | 19 | 32.8 |
| **Post Neoadjuvant therapy decision** | Surgery | 19 | 32.8 |
|  | Palliative treatment | 32 | 55.2 |
|  | Follow-up | 7 | 12 |
| **Post LCCRT was surgery done?** | Yes | 19 | 32.8 |
|  | No | 39 | 67.2 |
| **Type of Surgery done** | APR | 16 | 27.6 |
|  | LAR | 3 | 5.2 |
|  | Not done | 39 | 67.2 |

### Treatment Timelines

The interval from histologic diagnosis to initiation of radiotherapy ranged from 11 weeks to 153 weeks (Median: 64 weeks, IQR: 37–82). Among patients who underwent surgery, the median interval from completion of neoadjuvant therapy to surgery was 10 weeks (IQR, 7–15).

### Tumor Resectability

After completion of neoadjuvant therapy, tumors were reassessed for resectability by MDT. Overall, 27 patients (46.6%) were considered to have resectable disease. Of these, 19 ultimately underwent surgery. Reasons for deferring surgery among MDT-defined resectable patients included interval development of metastatic disease (n=11), patient refusal (n=4), and medical unfitness for surgery (n=4).

### Pathologic Response

Among the 19 patients who underwent surgery, pathologic assessment demonstrated no complete pathologic responses. Partial pathologic response of the primary tumor was observed in 6 patients (31.6%), while 11 patients showed no downstaging. For nodal disease, complete pathologic response was documented in 8 patients. (Table 3)

**Table 3:**
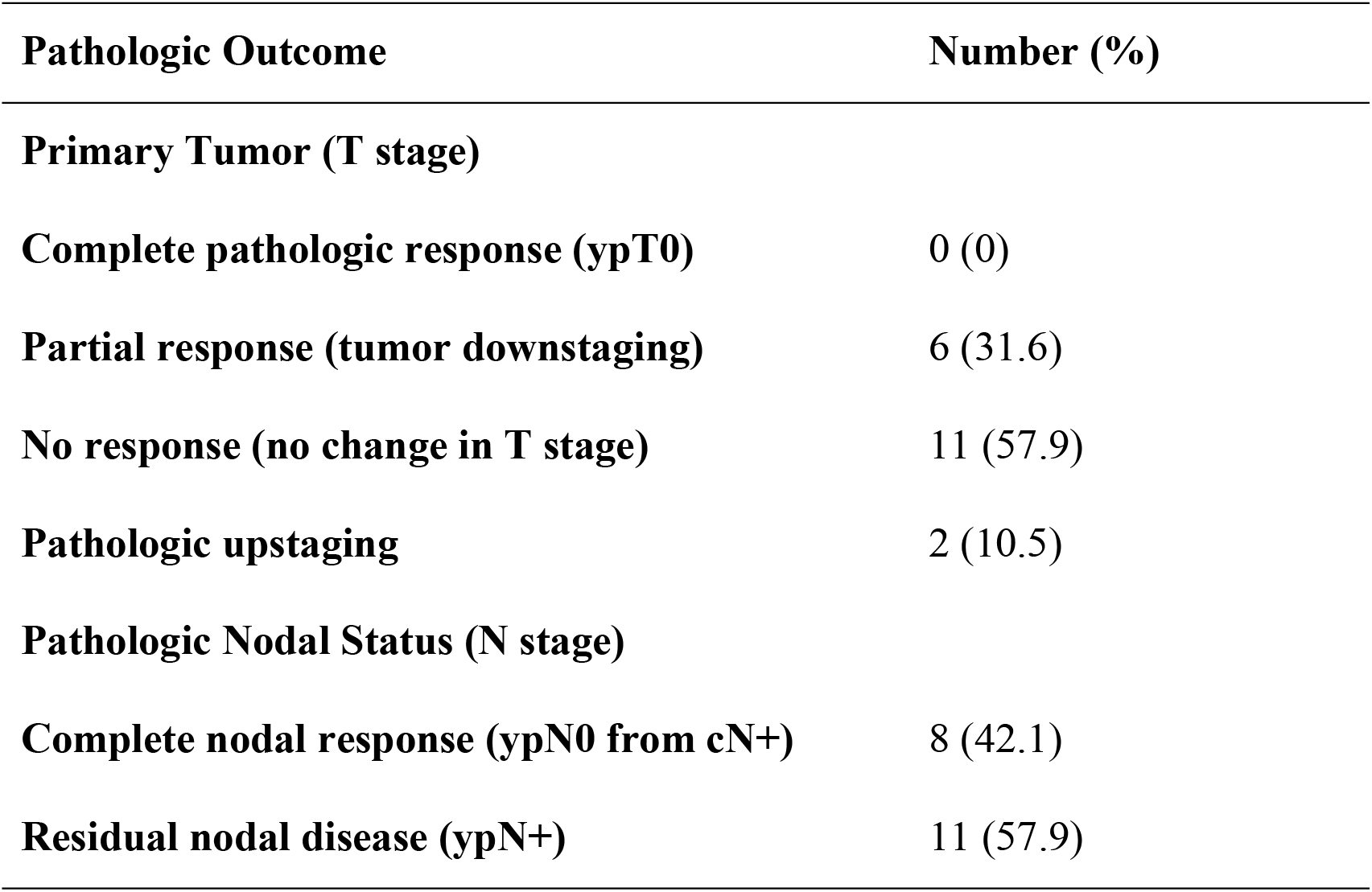
Pathologic Response in T stage in Locally Advanced Rectal Cancer patients at Tikur Anbessa Specialized Hospital, Ethiopia, October 2018 to October 2023 (n=19)

| **Pathologic Outcome** | **Number (%)** |
| --- | --- |
| **Primary Tumor (T stage)** |  |
| **Complete pathologic response (ypT0)** | 0 (0) |
| **Partial response (tumor downstaging)** | 6 (31.6) |
| **No response (no change in T stage)** | 11 (57.9) |
| **Pathologic upstaging** | 2 (10.5) |
| **Pathologic Nodal Status (N stage)** |  |
| **Complete nodal response (ypN0 from cN+)** | 8 (42.1) |
| **Residual nodal disease (ypN+)** | 11 (57.9) |

Negative proximal and distal margins were achieved in 15 patients (88.2%). Radial margins were negative in 11 patients (64.7%) and positive in 2 patients (11.8%). Lymphovascular invasion was reported in 8 patients, and perineural invasion in 4 patients.

### Factors associated with Tumor Resectability

Associations between baseline and treatment-related variables and MDT-defined tumor resectability were evaluated using univariable and multivariable logistic regression. Univariable analyses identified initial T stage, CRM status, neoadjuvant treatment strategy, histologic subtype, and radiotherapy technique as candidates for multivariable modeling.

In the multivariable model, only initial T stage and neoadjuvant treatment strategy remained significantly associated with resectability. Patients with cT3 disease had higher odds of resectability compared with those with cT4 disease (adjusted odds ratio [AOR], 6.21; 95% CI, 1.06–36.37). Patients treated with TNT were less likely to achieve resectability than those treated with LCCRT alone (AOR, 0.10; 95% CI, 0.02–0.49). Other variables, including CRM status and RT technique, were not independently associated with resectability after adjustment. Given the limited number of events, the model was restricted to key clinical variables to reduce overfitting; confidence intervals were wide, and results should be interpreted cautiously. (Table 4)

**Table 4:**
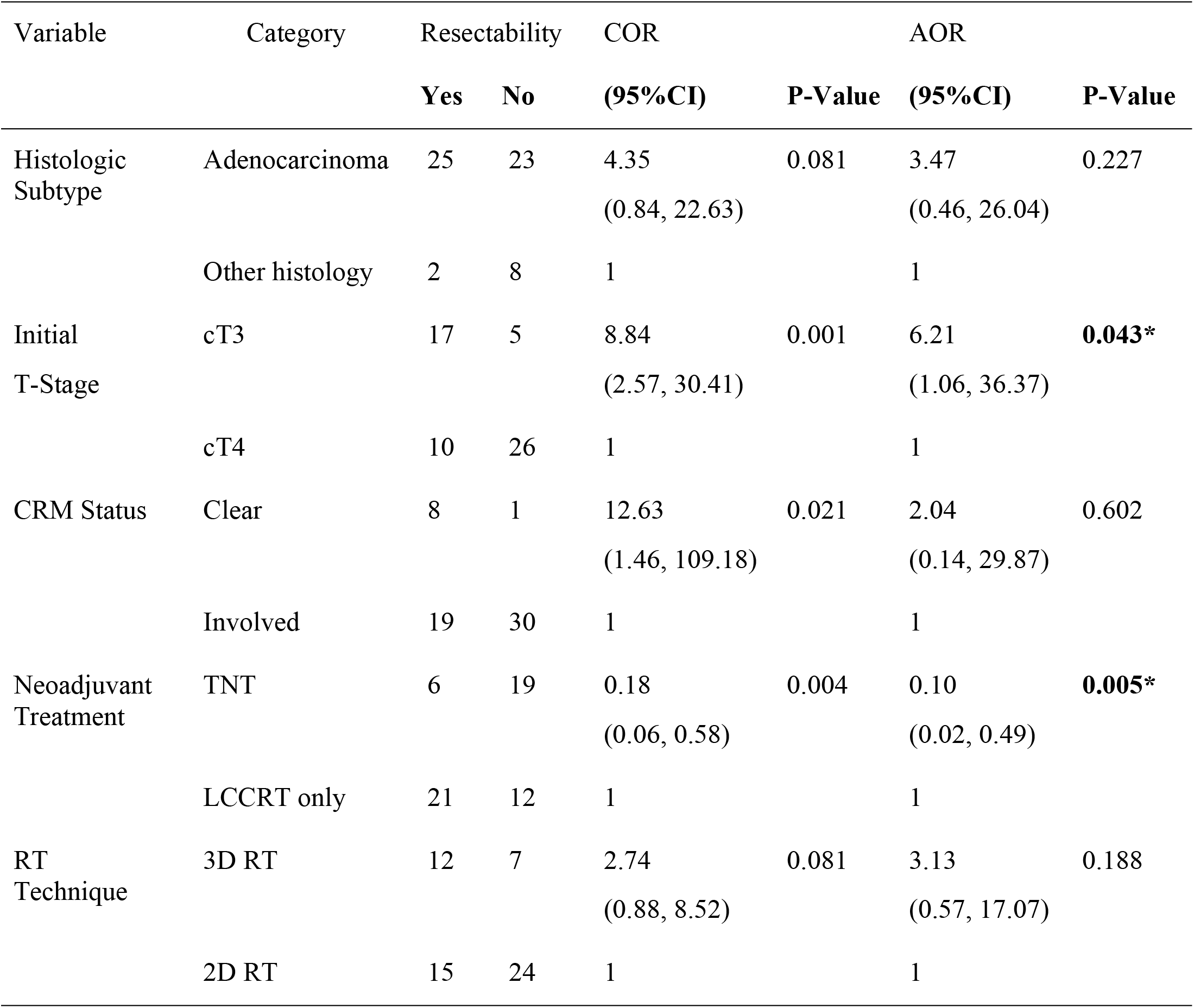
Factors Associated with Tumor Resectability after Neoadjuvant Treatment in Locally Advanced Rectal Cancer Patients at Tikur Anbessa Specialized Hospital, Ethiopia, October 2018 to October 2023. (N=58)

## Discussion

In this cohort of patients with LARC, tumor resectability and pathologic response following neoadjuvant therapy were lower than those reported in major contemporary trials (19, 20, 22, 26). These findings reflect a combination of advanced disease at presentation and prolonged delays across the treatment pathway, which are defining features of rectal cancer care in this resource-limited setting.

The demographic profile of the cohort differed from that reported in landmark trials evaluating LCCRT, SCRT, and TNT, in which the median age at diagnosis ranged from 55 to 67 years with male predominance (19, 20, 22, 26). In contrast, the median age in the current study was 45 years, with a slight female predominance. This observation is consistent with prior Ethiopian studies reporting a higher proportion of early-onset CRC and the younger demographics of the Ethiopian population (8, 23, 27, 28). Limited molecular data from the region suggest alterations in APC and KRAS genes and mismatch repair pathway changes among younger patients without a family history of CRC (27), raising the possibility that distinct tumor biology may contribute to the observed presentation and outcomes. However, molecular characterization was not available in this cohort and remains an important area for future investigation.

Disease burden at presentation was markedly advanced. In this study, 62% of patients had cT4 tumors, and the majority were initially deemed unresectable by the MDT assessment. While a similarly high proportion of cT4 disease was reported in the Polish II trial (65%) (21), the burden of advanced T stage in this cohort exceeded that seen in the RAPIDO and PRODIGE 23 trials, where cT4 disease accounted for approximately 30% and 16% of patients, respectively (19, 22). Nodal disease, although not typically a primary determinant of resectability, was also more advanced, with over half of patients presenting with cN2 disease, comparable to RAPIDO (22) but higher than PRODIGE 23 (19). These differences reveal that patients enrolled in randomized trials are often selected populations, whereas this cohort represents routine clinical practice with limited opportunities for early detection.

CRM involvement was another defining feature. In this cohort, CRM involvement on baseline pelvic MRI was reported in 84.5% of patients, higher than in RAPIDO (62%) and PRODIGE 23 (22%) (19, 22). Given the established association between CRM involvement and adverse oncologic outcomes, including reduced disease-free survival as demonstrated in the MERCURY study (29), the high prevalence of involved CRM likely contributed to both low resectability and limited pathologic response. This also explains the preference for LCCRT over SCRT in MDT decision-making, as downstaging and margin clearance are central objectives in patients with threatened CRM.

Although TNT is increasingly favored for LARC, especially in cT4 disease or tumors with mesorectal fascia involvement (12), its implementation in this setting was constrained by system-level factors. Despite 69% of patients meeting criteria for TNT, prolonged waiting times for radiotherapy limited its feasibility. Previous data from the same institution reported a median waiting time of more than 10 months from radiotherapy booking to treatment initiation (23), a finding mirrored in the current study, where the median interval from diagnosis to radiotherapy initiation was 62 weeks. Such delays likely allowed for disease progression and may have altered resectability between initial MDT assessment and treatment initiation, emphasizing the importance of reassessment with up-to-date imaging before radiotherapy.

Post-treatment tumor resectability in this cohort was markedly lower than that reported in large trials, where resectability rates exceeded 85% (19, 22). Multivariable analysis identified initial T stage as a key factor, with cT3 tumors significantly more likely to be deemed resectable than cT4 tumors. However, the wide confidence interval reflects limited statistical power and highlights the heterogeneity within cT4 disease, where outcomes vary depending on the extent and site of adjacent organ involvement. These findings reinforce the need for careful patient selection and realistic goal setting when neoadjuvant therapy is offered to patients with extensive locally advanced disease.

Patients treated with TNT were less likely to achieve resectability than those treated with LCCRT alone. This association likely reflects selection bias rather than treatment inefficacy, as patients assigned to TNT had a substantially higher burden of adverse prognostic features, including higher rates of cT4 disease and CRM involvement. Additionally, chemotherapy duration exceeded standard recommendations (12), with FOLFOX and CAPOX administered over extended periods, and unmeasured delays between treatment components may have further compromised outcomes. These deviations from trial protocols illustrate the challenges of translating intensive neoadjuvant strategies into settings with limited capacity.

Radiotherapy Technique of delivery is not commonly mentioned in the literature as a factor that contributes to oncologic outcomes in rectal cancer, even though highly conformal techniques like Intensity Modulated Radiotherapy (IMRT) have been associated with less radiation toxicity (31). In this study, most patients were treated using 2D RT with a Cobalt-60 Radiotherapy machine, as the study period included the time before a Linear Accelerator Radiotherapy machine was installed at TASH. Although all patients completed planned radiotherapy, the impact of prolonged overall treatment time and limited conformality on tumor response cannot be excluded.

The quality of life of rectal cancer survivors is an important issue to consider. In this regard, sphincter-preserving surgeries are preferred to APR, which requires a permanent colostomy. APR was performed in the majority of patients, consistent with the high prevalence of low-rectal tumors. While negative proximal and distal margins were commonly achieved, the radial margin was reported as negative in 64.7 %, which is lower compared to R0 resection in RAPIDO with 90 % and PRODIGE 23 with 94 % (19, 22).

Pathologic response was limited, with no pathologic complete responses observed, and more than half of the patients showed no T-stage downstaging. This contrasts with reports from neighboring regions, including a Sudanese study that demonstrated higher rates of downstaging and pathologic complete response after LCCRT (34). Delayed surgery may have contributed to this finding, as the median interval between completion of LCCRT and surgery was 10 weeks, exceeding commonly recommended intervals of 6–8 weeks (17, 19, 30).

A clinical complete response is currently being considered as a surrogate marker for pathologic complete response rates, but none of the patients achieved this degree of response. Meta-analyses reported that having a complete pathologic response rendered a better prognosis in rectal cancer patients than a partial or no pathologic response. There was a local control, DFS, and overall survival advantage (32, 33). The limited tumor regression observed in this cohort is likely to translate into inferior long-term outcomes, as suggested by prior Ethiopian studies (8, 28). Distinct patient profiles and tumor molecular biology could have contributed to the poor pathologic response in this study, which needs further investigation.

Overall, these findings emphasize that advanced disease at presentation, prolonged treatment delays, and limitations in radiotherapy and surgical capacity collectively shape rectal cancer outcomes in this setting. Addressing these structural constraints is central to improving the effectiveness of neoadjuvant strategies in routine clinical practice.

## Conclusion

Most patients in this cohort presented with advanced rectal cancer characterized by cT4 disease and CRM involvement, and more than half remained unresectable despite completion of neoadjuvant therapy. Tumors that were ultimately deemed resectable were predominantly cT3, and pathologic response was limited, with no complete responses observed. Prolonged delays in radiotherapy initiation and surgery likely contributed to disease progression and reduced curative potential. Therefore, careful patient selection with MDT and timely administration of planned radiotherapy are mandatory to ensure that patients benefit from neoadjuvant treatment. Further studies incorporating molecular characterization of rectal cancer in the Ethiopian population may help clarify biologic contributors to treatment response and guide future therapeutic strategies.

## Limitation of the study

Some methodological constraints should be considered when interpreting these findings. The retrospective design limited data completeness and resulted in a relatively small sample size, reducing statistical power and contributing to wide confidence intervals in multivariable analyses. Tumor resectability was defined by MDT assessment rather than standardized trial-based criteria, which may limit direct comparability with prospective studies. In addition, restaging imaging was not routinely performed immediately before initiation of chemoradiotherapy, raising the possibility of stage migration during prolonged treatment intervals. Finally, as a single-center experience, the findings may not be broadly generalizable but likely reflect challenges common to similar resource-limited oncology settings.

## Data Availability

All relevant data are contained within the manuscript and supporting information files.

## Acknowledgments

We wish to thank the staff at the Department of Clinical Oncology, Tikur Anbessa Specialized Hospital, for their profound support.

## Author Contributions

**Samrawit Seyoum Halake:** Conceptualization (lead), methodology (lead), data curation (lead), formal analysis (equal), investigation (lead), funding acquisition (equal), resources (lead), validation (equal), writing – original manuscript (lead); writing – review and editing (equal).

**Hawi Furgassa Bedada:** Conceptualization (supporting), methodology (supporting), data curation (supporting), formal analysis (equal), investigation (supporting), validation (equal), writing – original manuscript (equal), writing – review and editing (equal).

**Tariku Mengesha Desalegn:** Methodology (supporting), formal analysis (equal), investigation (supporting), validation (equal), writing – original manuscript (supporting), writing – review and editing (equal).

**Tsige Beyene Feyisa:** Methodology (supporting), data curation (supporting), validation (supporting), writing – original manuscript (supporting), writing – review and editing (equal).

**Kibrekidusan Aynekulu Tsige:** Methodology (supporting), data curation (supporting), validation (supporting), writing – original manuscript (supporting), writing – review and editing (equal).

**Edom Seife Woldetsadik:** Supervision (lead), methodology (supporting), funding acquisition (equal), resources (supporting), validation (equal), writing – original manuscript (supporting), writing – review and editing (equal).

**Eva Johanna Kantelhardt:** Supervision (supporting), methodology (supporting), funding acquisition (lead), resources (supporting), validation (equal), writing – original manuscript (supporting), writing – review and editing (equal).

